# Psychological trauma and the genetic overlap between posttraumatic stress disorder and major depressive disorder

**DOI:** 10.1101/2020.11.25.20229757

**Authors:** Jessica Mundy, Christopher Hübel, Joel Gelernter, Daniel Levey, Robin M. Murray, Megan Skelton, Murray B. Stein, The Million Veteran Program, Post Traumatic Stress Disorder Working Group of the Psychiatric Genomics Consortium, Evangelos Vassos, Gerome Breen, Jonathan R. I. Coleman

**Affiliations:** Social, Genetic and Developmental Psychiatry Centre; Institute of Psychiatry, Psychology & Neuroscience, King’s College London, London, United Kingdom; UK National Institute for Health Research (NIHR) Biomedical Research Centre, South London and Maudsley National Health Service (NHS) Trust, London, United Kingdom; Department of Medical Epidemiology and Biostatistics, Karolinska Institutet, Stockholm, Sweden; Institute of Psychiatry, Psychology & Neuroscience, King’s College London, London, United Kingdom; Division of Human Genetics, Department of Psychiatry, Yale University School of Medicine, New Haven, Connecticut, United States of America; Department of Psychiatry, Veterans Affairs Connecticut Healthcare Center, West Haven, Connecticut, United States of America; Departments of Genetics and Neuroscience, Yale University School of Medicine, New Haven, Connecticut, United States of America; Psychiatry Service, VA San Diego Healthcare System, San Diego, California, United States of America; Departments of Psychiatry and Family Medicine & Public Health, University of California San Diego, La Jolla, California, United States of America

## Abstract

**Background:** Posttraumatic stress disorder (PTSD) and major depressive disorder (MDD) are commonly reported co-occurring mental health consequences following psychological trauma exposure. The disorders have high genetic overlap. We investigated whether the genetics of PTSD were associated with reported trauma in individuals with MDD. Since trauma is associated with recurrent MDD, we also investigated whether the genetics of PTSD were associated with episode recurrence.

**Methods:** Genetic correlations were estimated between PTSD and MDD in the presence and MDD in the absence of reported exposure to psychological trauma, and recurrent and single-episode MDD, based on genetic data from UK Biobank Mental Health Questionnaire respondents (*N*=157,358). Genetic correlations were replicated using PTSD data from the Psychiatric Genomics Consortium and Million Veteran Program. Polygenic risk scores were generated to investigate whether individuals with MDD who have higher genetic risk for PTSD were more likely to report psychological trauma than those with lower genetic risk.

**Results:** Individuals with MDD with a higher genetic risk for PTSD were significantly more likely to report exposure to psychological trauma than those with lower risk [OR=1.06 (1.03-1.09) Empricial *p*<0.001]. PTSD was significantly more genetically correlated with recurrent MDD than with MDD in the absence of reported psychological trauma [*r*_g_ differences = ∼0.2, *p*<0.008]. Participants who had experienced recurrent depressive episodes reported significantly higher trauma rates than participants who had experienced a single episode [*chisquare*>167, *p*<0.001].

**Conclusions:** Genetic risk for PTSD in individuals with MDD may influence the way in which traumatic life events are perceived, responded to and reported.

## Introduction

Symptoms of posttraumatic stress disorder (PTSD) and major depressive disorder (MDD) are the most commonly described co-occuring problems following exposure to psychological trauma (Ben Barnes *et al*., 2018). Across epidemiological samples, approximately 50% of individuals with PTSD have a comorbid diagnosis of MDD (Kessler *et al*., 1995; Breslau *et al*., 1997; Rytwinski *et al*., 2013). Similar, or occasionally higher, estimates are observed in primary care settings (Stein *et al*., 2000; Alim *et al*., 2006). Previously, high comorbidity rates have been attributed to the classification of shared symptoms into the two diagnostic categories (Flory and Yehuda, 2015), such as negative mood, sleep disturbances, irritability, and concentration difficulties (American Psychiatric Association, 2013). However, several studies demonstrate that the ∼50% comorbidity rate does not diminish after excluding these shared symptoms from clinical diagnoses (Grubaugh *et al*., 2010; Elhai *et al*., 2011) suggesting the observed symptom overlap does not provide an adequate explanation. An alternative explanation might be genetic overlap between the disorders. Twin studies have previously indicated that PTSD shares genetic influences with MDD (*r*=0.77) and related conditions (Koenen *et al*., 2008; Wolf *et al*., 2010). More recently, methods based on genome-wide association studies (GWAS) have been used to explore genetic correlations (*r*_g_), a quantitative measure of the genetic relationship between two polygenic traits (van Rheenen *et al*., 2019). Research from the Psychiatric Genomics Consortium (PGC) reported strong, positive genetic correlations of PTSD with depressive symptoms (*r*_g_=0.80) and with MDD (*r*_g_=0.62) (Nievergelt *et al*., 2019), thus supporting results from twin studies. As well as shared genetics, another potential factor involved in PTSD-MDD comorbidity is exposure to trauma. There is a complex relationship between trauma exposure and mental health sequelae. Exposure to trauma is common (Breslau *et al*., 1991; Kessler *et al*., 1995). 50-90% of people will experience a traumatic event in their lifetime but only 8-12% will go on to develop PTSD (Shah *et al*., 2012), suggesting that certain individuals are at greater risk of developing PTSD than others following exposure (Auxéméry, 2012; Duncan *et al*., 2018b; Nievergelt *et al*., 2019). Similarly, stressful and traumatic events are significant risk factors for MDD (Horesh *et al*., 2008; Shapero *et al*., 2014; Hovens, 2015), but the majority of people who are exposed do not develop the disorder (Kessler, 1997). Therefore, similar to PTSD risk, the effects of trauma on MDD risk are moderated by individual liability (Colodro-Conde *et al*., 2018). Previous research has shown that reported traumatic events are not only heritable themselves (Jay Schulz-Heik *et al*., 2009; Power *et al*., 2013; Dalvie *et al*., 2020) but also increase the SNP-based heritability of MDD (Coleman *et al*., 2020), demonstrating that genetic factors influence not only on the risk of both being exposed, but also on the risk of developing a mental health disorder following exposure.

### Aims

The ∼50% PTSD-MDD comorbidity rate demonstrates that not everyone responds to psychological trauma in the same way. This may be due to genetic liability in trauma sensitivity. No study has investigated whether the extent to which PTSD and MDD overlap genetically is associated with exposure to psychological trauma. We addressed this by examining genetic correlations between PTSD and four MDD phenotypes in the UK Biobank, with replication using the largest PTSD GWASs to data from the PGC and the Million Veteran Program (MVP). Given evidence from clinical studies, we hypothesized that PTSD and MDD with reported trauma would have higher genetic overlap compared to PTSD and MDD without reported trauma, which would add further evidence for the existence of genetic variants associated with trauma sensitivity. We also explored the genetic overlap of PTSD with single-episode MDD and with recurrent MDD. Research has shown that the type, frequency, and severity of traumatic events are associated with the frequency and severity of subsequent depressive episodes (Nanni *et al*., 2012; Hovens *et al*., 2015; Otte *et al*., 2016), with childhood maltreatment being particularly associated with MDD recurrence (Danese, 2020). Accordingly, our second hypothesis was that PTSD would have greater genetic overlap with recurrent MDD compared to single-episode MDD, under the assumption that rates of trauma are higher among individuals with recurrent MDD. We tested the validity of this assumption in UK Biobank participants with MDD. To further address our research question, we generated PTSD polygenic risk scores (PRS) in UK Biobank participants with MDD to examine whether there is an association between genetic risk for PTSD and the reporting of traumatic events. Following the logic of our previous hypotheses, we expected individuals with MDD with higher genetic risk for PTSD would be more likely to report experiencing psychological trauma and would be more likely to have experienced recurrent depressive episodes than those with lower genetic risk for PTSD.

## Methods

This study used summary statistics from previous GWAS of 1) self-reported PTSD in UK Biobank (Nievergelt *et al*., 2019), 2) MDD with reported psychological trauma exposure, 3) MDD without reported psychological trauma exposure (Coleman *et al*., 2020), 4) recurrent MDD, and 5) single-episode MDD (Coleman *et al*., 2019). All phenotypes were based on UK Biobank participants who responded to the follow-up online Mental Health Questionnaire (*N*=157,358). PTSD phenotypes can reflect the characteristics of the sample and data collection method. To examine whether our findings were consistent across differing PTSD phenotypes, we repeated the genetic correlations using additional sets of summary statistics (Table1). We used results from a GWAS of mainly clinical samples of PTSD undertaken by the PGC, known as PGC 1.5 PTSD (Nievergelt *et al*., 2019). We also repeated our analyses using the meta-analysis of PGC 1.5 and UK Biobank PTSD, known as PGC 2 PTSD (Nievergelt *et al*., 2019). Finally, we used a set of summary statistics from a GWAS of United States veterans by the MVP, based on electronic health records (Stein *et al*., 2020).

**Table 1:**
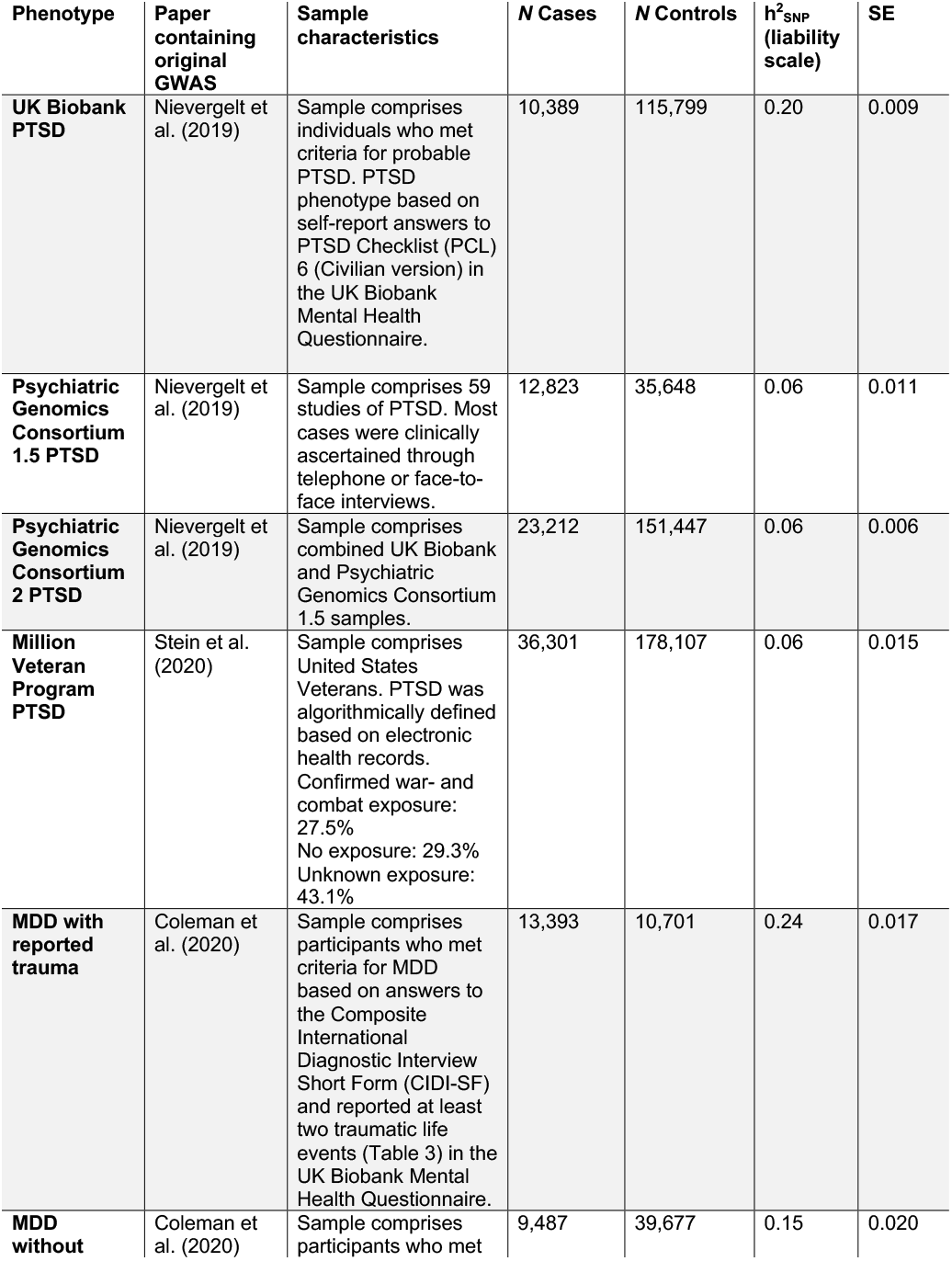
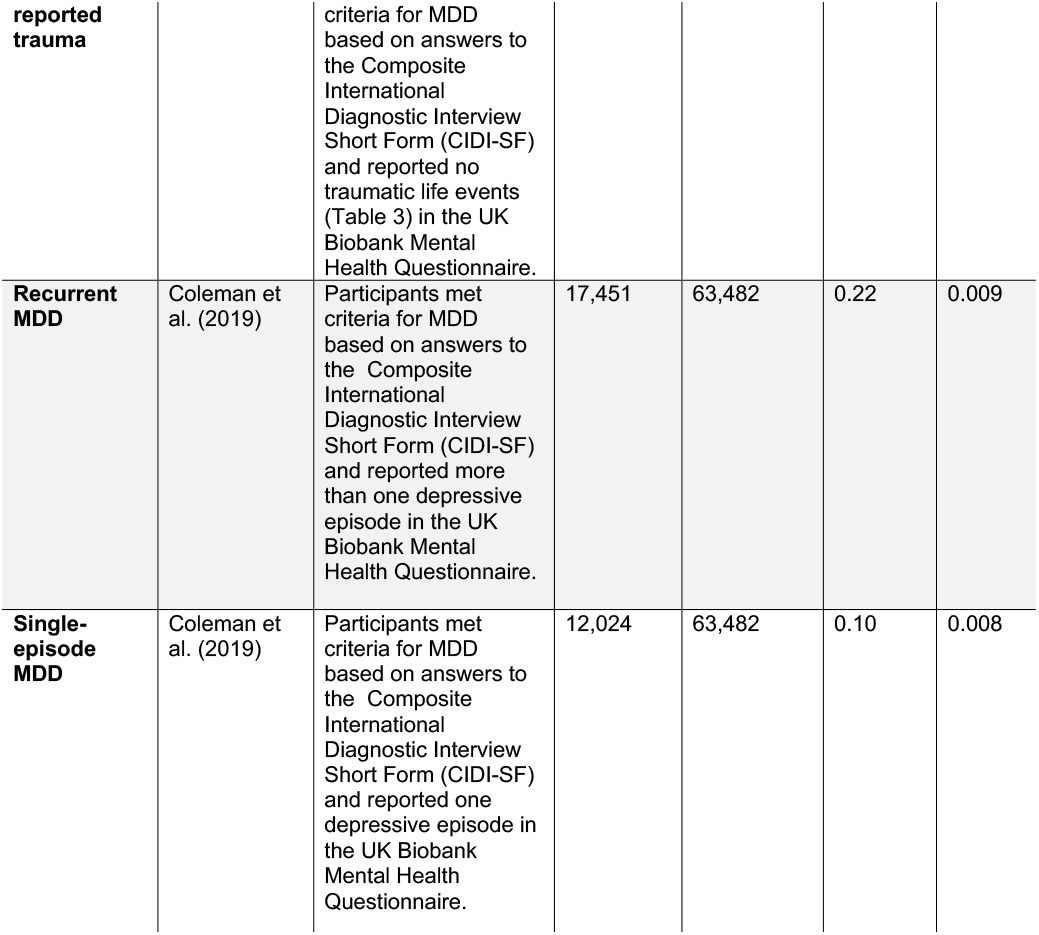
Information about the four posttraumatic stress disorder (PTSD) and four major depressive disorder (MDD) genome-wide association study (GWAS) summary statistics, including the original publication, characteristics of the sample, number (*N*) of cases and controls in original GWAS, liability scale SNP-based heritability (h^2^_SNP_) and standard error (SE) from High Definition Likelihood. Details of how observed scale h^2^_SNP_ estimates were converted to the liability scale are presented in the Supplementary Methods.

Using the MVP PTSD summary statistics, we generated PRS for participants who met criteria for MDD in the UK Biobank. The number of cases and controls and the SNP-based heritability (on the liability scale) of each GWAS can be found in Table 1. All summary statistics were produced from GWAS on individuals of European ancestries. Details of the contributing studies and phenotype definitions can be found in the Supplementary Methods. Brief details are presented in Table 1.

### Reported trauma exposure in the individuals with MDD in the UK Biobank

We tested the assumption behind our second hypothesis, which states that the rates of trauma exposure are higher among individuals who experience recurrent as opposed to single-episode MDD. In the UK Biobank, participants were categorised as having experienced either recurrent or single-episode MDD, as defined by Coleman et al. (2019). For seven traumatic life events, which were included in the Coleman et al. (2020) definition of “reported trauma exposure” (Table 3), we compared reporting rates between those in who met criteria for recurrent MDD and those who met criteria for single-episode MDD. We performed chi-square tests in R to establish whether there were differences in trauma reporting rates. Chi-square tests were considered statistically significant if they reached or surpassed the Bonferroni-corrected alpha (0.05/7=0.007; i.e. to correct for the seven chi-square tests performed).

Throughout this paper, any mention of trauma exposure in participants from the UK Biobank refers specifically to retrospective self-reported psychologically traumatic events due to the nature of data collection via an online questionnaire. The events being reported may have occurred before, after, or concurrently with MDD episodes (Coleman *et al*., 2020).

### Genetic correlations

GWAS summary statistics (Table 1) were used to calculate genetic correlations based on single nucleotide polymorphisms (SNP-based *r*_g_) using the High Definition Likelihood (HDL) software and the 1,029,876 quality controlled UK Biobank imputed HapMap3 SNPs reference panel. This reference panel is based on genotypes in UK Biobank, which were imputed to HRC and UK10K + 1000 Genomes (Ning *et al*., 2020).

First, we calculated genetic correlations between PTSD and (i) MDD with reported trauma, (ii) MDD without reported trauma, (iii) recurrent MDD, and (iv) single-episode MDD within the UK Biobank. We then replicated these genetic correlations using the PGC 1.5 PTSD phenotype, the combined PGC 2 phenotype and the MVP phenotype. Genetic correlations were tested for a significant difference from 0 (default in HDL) and for a difference from 1 (in Microsoft Excel, converting *r*_g_ to a chi-square as [(*r*_g_ − 1)/ se]^2^). An explanation of HDL inference of genetic correlations can be found in Ning et al. (2020). Genetic correlations were considered significantly different to 0 or to 1 if they surpassed the Bonferroni-corrected alpha in each analysis (0.05/4=0.0125; i.e. to correct for the four tests in each independent set of correlations).

To test the significance of the differences between the genetic correlations we performed a block-jackknife, which uses resampling to recalculate standard errors for the differences between two *r*_g_ estimates. Within each of the four groups of correlations (i.e. for the four different PTSD phenotypes) we compared *r*_g_ estimates in a pairwise fashion (i.e. each correlation pair was compared with all other pairs within the group). This resulted in six different block-jackknife tests per PTSD phenotype. Differences between genetic correlations were considered statistically significant if they surpassed the Bonferroni corrected alpha (0.05/6=0.0083; i.e. to correct for the six tests).

The Supplementary Methods contains further details of the HDL analysis, including percentage overlap between the summary statistics and the HapMap3 reference panel.

We also ran these analyses using Linkage Disequilibrium Score Regression (LDSC), another command line tool for estimating heritability and genetic correlations from GWAS summary statistics (Bulik-Sullivan *et al*., 2015). In our study, we favoured HDL for estimating genetic correlations. Unlike LDSC, HDL uses a full likelihood-based method to estimate genetic correlations that fully accounts for linkage disequilibrium (LD) across the genome. When compared to LDSC, HDL reduces the variance of the genetic correlation by approximately 60% (Ning *et al*., 2020). Consequently, HDL is better powered to detect significant differences between correlations, which was a central aim of our study. The LDSC results and an explanation of any differences from HDL are presented in the Supplementary Results.

### Polygenic risk scores

We calculated individual PRS for PTSD using the MVP summary statistics in individuals with MDD from the UK Biobank. For this, we used PRSice v2.3.1 and controlled for the first six principal components, genotyping batch and assessment centre. PRS were calculated at 11 *p*-value thresholds (5×10^−8^, 1×10^−5^, 1×10^−3^, 0.01, 0.05, 0.1, 0.2, 0.3, 0.4, 0.5, 1). Phenotype permutations were used to produce an empirical *p*-value for the association at the best-fitting PRS, which accounts for testing at multiple thresholds (Euesden *et al*., 2015). Once the best-fitting PRS had been calculated, we then performed logistic regressions to examine whether genetic risk for PTSD was more strongly associated with MDD with reported trauma compared to MDD without reported trauma, and to examine whether genetic risk for PTSD was more strongly associated with recurrent compared to single-episode MDD. The standardised beta coefficients were converted to odds ratios (OR) in R and 95% confidence intervals (CI) were calculated. The full six pairwise comparisons, as in the block-jackknife analysis in this study, were not possible due to MDD cases in the UK Biobank belonging to overlapping MDD subtypes. Therefore, we limit the PRS analysis to these two comparisons.

We performed power calculations using the Additive Variance Explained and Number of Genetic Effects Method of Estimation (AVENGEME) programme in R (Dudbridge, 2013). Details of this are presented in the Supplementary Methods. The MVP summary statistics were chosen due to their power and there being no overlap between the individuals in this sample and the target sample (UK Biobank). Since one target sample was used to produce PRS for the two regressions a Bonferroni adjustment was used correcting for 2 tests and giving a final threshold of *p*<0.025.

## Results

### Trauma exposure in the UK Biobank

Seven traumatic life events comprised the overall definition of “trauma exposure” in Coleman et al. (2020) (Table 3). Individuals who reported two or more events were considered “trauma-exposed”. Each of the seven life events were significantly more commonly reported by UK Biobank participants who reported having experienced recurrent MDD than those who had experienced single-episode MDD. This confirms our assumption that the group of participants in the UK Biobank with recurrent MDD demonstrate a higher rate of psychological trauma exposure than those in the single-episode group.

### Genetic correlations

All genetic correlations were significantly different from 0. The genetic correlations between both PGC 1.5 and PGC 2 PTSD and single-episode MDD were found to not differ significantly from 1, although this finding is likely due to the large standard errors of the *r*_g_ estimates, reflecting low power. All other genetic correlations were significantly different to one (Table 4).

**Figure 1:**
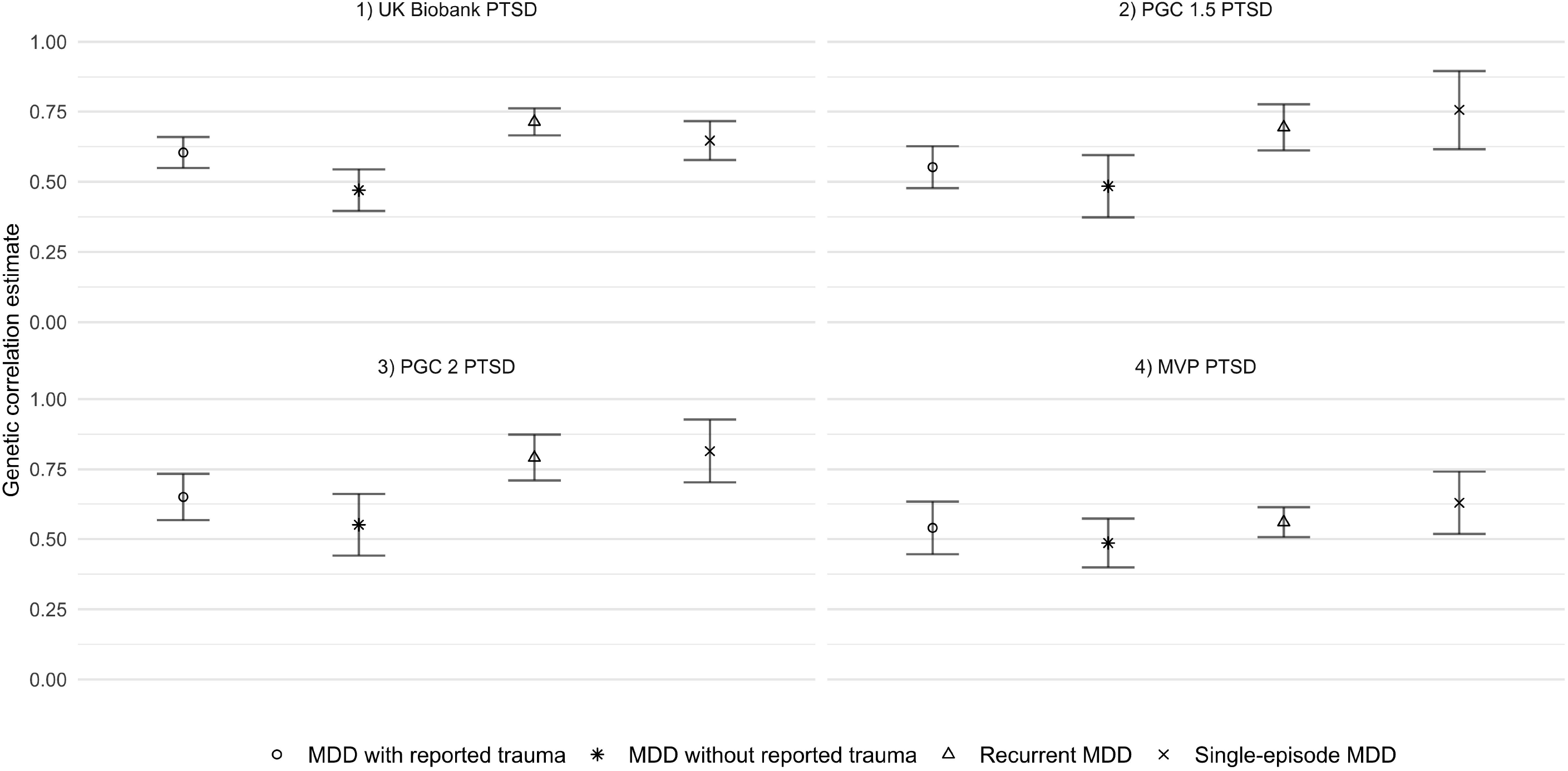
High Definition Likelihood (HDL) genetic correlation (*r*_*g*_) estimates of four PTSD phenotypes 1) UK Biobank posttraumatic stress disorder (PTSD), 2) Psychiatric Genomics Consortium (PGC) PTSD 1.5, 3) PGC PTSD 2 and 4) Million Veteran Program (MVP) PTSD with the four major depressive disorder (MDD) phenotypes. Standard errors are shown in the error bars surrounding the *r*_*g*_ estimates for each genetic correlation.

### Differences between genetic correlations of PTSD and MDD phenotypes

The genetic correlation between PTSD and recurrent MDD was significantly greater than that between PTSD and MDD without reported trauma when using the UK Biobank, PGC 1.5 and PGC 2 phenotypes. All other genetic correlations were not significantly different from each other (Supplementary Table 1). Genetic correlation estimates of PTSD with MDD with reported trauma were consistently larger than those with MDD without reported trauma, albeit not significant (*p*=0.14 – 0.65) (Table 4, Supplementary Table 1). In contrast, no consistent pattern of genetic correlation was observed between PTSD and recurrent versus single-episode MDD (Table 3, Supplementary Table 1).

**Table 2:** Difference in reporting rates of traumatic life events between individuals with recurrent and single-episode major depressive disorder (MDD) in UK Biobank Mental Health Questionnaire Respondents (*N*=157,358). Differences were considered significant if they surpassed the Bonferroni adjusted alpha (*p*<0.007)

| Trauma category | Traumatic event | Endorsement in single-episode MDD (%) | Endorsement in recurrent MDD (%) | $\chi^2$ statistic | P-value |
| --- | --- | --- | --- | --- | --- |
| Childhood emotional abuse | Felt hated by a family member as a child | 2,352 (8%) | 5,239 (18%) | 405 | $3.52 \times 10^{-16}$ |
| Childhood emotional neglect | Did not feel loved as a child | 3,122 (11%) | 6,702 (23%) | 497 | $3.73 \times 10^{-110}$ |
| Childhood sexual abuse | Was sexually molested as a child | 1,217 (4%) | 2,690 (9%) | 175 | $6.39 \times 10^{-16}$ |
| Adulthood emotional abuse | Was belittled by a partner or ex-partner | 3,887 (13%) | 7,591 (26%) | 371 | $1.27 \times 10^{-82}$ |
| Adulthood physical abuse | Was physically abused by a partner or ex-partner | 2,005 (7%) | 3,987 (14%) | 167 | $4.29 \times 10^{-38}$ |
| Adulthood sexual abuse | Was forced to have sex against my will by a partner or ex-partner | 890 (3%) | 2,283 (8%) | 240 | $3.26 \times 10^{-54}$ |
| Sexual assault | Ever been a victim of sexual assault | 2,247 (8%) | 4,764 (16%) | 292 | $1.05 \times 10^{-65}$ |

**Table 3:** High Definition Likelihood genetic correlation estimates (*r*_g_) and standard errors (SE) of 1) UK Biobank posttraumatic stress disorder (PTSD), 2) Psychiatric Genomics Consortium (PGC) 1.5 PTSD, (3 PGC 2 PTSD and 4) Million Veteran Program (MVP) PTSD with the four major depressive disorder (MDD) phenotypes. *P* (diff 0) refers to *p*-value for test of *r*_g_ different from 0. *P* (diff 1) refers to *p*-value for test of *r*_g_ different from 1. Genetic correlations were considered significant if they surpassed the Bonferroni adjusted threshold (*p*<0.0125).

| PTSD Phenotype | MDD phenotype | $r_g$ | SE | $P$ (diff 0) | $P$ (diff 1) |
| --- | --- | --- | --- | --- | --- |
| UK Biobank PTSD | MDD with reported trauma | 0.6040 | 0.06 | $4.92 \times 10^{-28}$ | $6.02 \times 10^{-13}$ |
| UK Biobank PTSD | MDD without reported trauma | 0.4701 | 0.07 | $2.43 \times 10^{-10}$ | $9.23 \times 10^{-13}$ |
| UK Biobank PTSD | Recurrent MDD | 0.7134 | 0.05 | $1.03 \times 10^{-49}$ | $2.55 \times 10^{-9}$ |
| UK Biobank PTSD | Single-episode MDD | 0.6466 | 0.07 | $8.35 \times 10^{-21}$ | $3.15 \times 10^{-7}$ |
| PGC 1.5 PTSD | MDD with reported trauma | 0.5520 | 0.07 | $1.35 \times 10^{-13}$ | $1.91 \times 10^{-9}$ |
| PGC 1.5 PTSD | MDD without reported trauma | 0.4841 | 0.11 | $1.22 \times 10^{-5}$ | $3.16 \times 10^{-6}$ |
| PGC 1.5 PTSD | Recurrent MDD | 0.6937 | 0.08 | $2.94 \times 10^{-17}$ | $1.91 \times 10^{-4}$ |
| PGC 1.5 PTSD | Single-episode MDD | 0.7560 | 0.14 | $7.09 \times 10^{-8}$ | 0.08 |
| PGC 2 PTSD | MDD with reported trauma | 0.6497 | 0.08 | $3.39 \times 10^{-15}$ | $2.18 \times 10^{-5}$ |
| PGC 2 PTSD | MDD without reported trauma | 0.5509 | 0.11 | $5.12 \times 10^{-7}$ | $4.24 \times 10^{-5}$ |
| PGC 2 PTSD | Recurrent MDD | 0.7915 | 0.08 | $8.45 \times 10^{-22}$ | 0.01 |
| PGC 2 PTSD | Single-episode MDD | 0.8147 | 0.11 | $5.27 \times 10^{-13}$ | 0.1 |
| MVP PTSD | MDD with reported trauma | 0.5397 | 0.09 | $8.77 \times 10^{-9}$ | $9.24 \times 10^{-7}$ |
| MVP PTSD | MDD without reported trauma | 0.4859 | 0.09 | $2.41 \times 10^{-8}$ | $3.58 \times 10^{-9}$ |
| MVP PTSD | Recurrent MDD | 0.5600 | 0.05 | $6.57 \times 10^{-26}$ | $1.33 \times 10^{-16}$ |
| MVP PTSD | Single-episode MDD | 0.6291 | 0.11 | $1.59 \times 10^{-8}$ | $8.61 \times 10^{-4}$ |

### Polygenic risk scores

In individuals with MDD in the UK Biobank, those with a higher genetic risk for PTSD were significantly more likely to report trauma than those with a lower PTSD risk (OR=1.06 (1.03-1.09) Empirical *p*<0.001; Table 5). In contrast, those with a higher genetic risk for PTSD were more likely to have experienced recurrent episodes but this was not significant (OR=1.02 (1.00-1.05) Empirical *p*=0.28; Table 5). The variance explained by the PRS was low, ranging from 0.02% to 0.13% based on varying the assumed population prevalence of the target phenotype. See Supplementary Results for full details of this analysis, including the number of SNPs in each PRS and the Nagelkerke’s R^2^ for a range of population prevalence (Supplementary Table 4).

**Table 5:** Results of posttraumatic stress disorder (PTSD) polygenic risk score (PRS) regression analysis on individuals in the UK Biobank with major depressive disorder (MDD), including the odds ratio (OR) and 95% confidence intervals (CI), *p*-value (*P*) and empirical *p*-value accounting for testing at multiple thresholds (Empirical *P*). OR were considered significant if the Empirical *P* surpassed the Bonferroni adjusted threshold (Empirical *p*-value<0.025).

| Regression | OR (95% CI) | *P* | Empirical *P* |
| --- | --- | --- | --- |
| **MDD with reported trauma vs. MDD without reported trauma** | 1.06 (1.03 - 1.09) | 2.11X10<sup>-5</sup> | 0.001 |
| **Recurrent MDD vs. single-episode MDD** | 1.02 (1.00 - 1.05) | 0.05 | 0.28 |

## Discussion

This study investigated whether the genetics of PTSD were more strongly overlapping with the genetics of MDD with reported trauma compared to MDD without reported trauma. This was based on clinical observations of high comorbidity among individuals who had been exposed to traumatic events, and evidence from genomic studies that the disorders strongly overlap in terms of additive genetic variants. We also investigated whether genetic risk for PTSD was associated with the risk of reporting trauma exposure, a trait known to be heritable, in UK Biobank participants with MDD. Across multiple PTSD GWAS, the difference in genetic correlation between PTSD and MDD with reported trauma and MDD without reported trauma was not significant. This indicates that any true difference is not large – however, we were underpowered to detect small differences in genetic correlation which means we cannot draw strong conclusions from this analysis alone. By contrast, the findings from the PRS analysis demonstrated that individuals with MDD with a higher genetic risk for PTSD were significantly more likely to report traumatic life events than those who had a lower genetic risk for PTSD.

In addition to these findings, we note that UK Biobank participants who met criteria for recurrent MDD reported significantly higher rates of trauma exposure in comparison to individuals who met criteria for single-episode MDD. This corroborates previous psychiatric research which pinpoints exposure to stressful or traumatic events as a key risk factor for subsequent recurrent MDD (Nanni *et al*., 2012; Hovens *et al*., 2015; Otte *et al*., 2016). Based upon this, we expected PTSD to show a greater genetic correlation with recurrent MDD phenotype compared to the single-episode MDD phenotype. However, we found no evidence of this in the genetic correlation analysis. Furthermore, findings from the PRS analysis show that genetic risk for PTSD was not more strongly associated with recurrent compared to single-episode MDD.

Previous research has postulated that PTSD-MDD comorbidity represents a specific trauma-related psychiatric trait (Flory and Yehuda, 2015), perhaps indicating a sensitivity to traumatic or stressful events. In our study, we used UK Biobank data on traumatic life events which participants self-reported via the online Mental Health Questionnaire. Previous research from twin studies has shown that the reporting of trauma has a heritable basis (Jay Schulz-Heik *et al*., 2009). More recently, genomic studies have suggested that at least part of this heritability can be attributed to additive genetic variants, reporting a SNP-based heritability estimate at around 18% for lifetime trauma (Coleman *et al*., 2020), and around 6% specifically for childhood trauma (Dalvie *et al*., 2020). The findings in our study from the PRS analysis suggest that genetic liability for PTSD is associated with the reporting of psychological trauma in those who have MDD. Experiencing trauma is common, but only a minority of individuals develop PTSD (Auxéméry, 2012; Duncan *et al*., 2018a; Nievergelt *et al*., 2019). The PTSD summary statistics used to generate PRS in this study are therefore capturing these individual differences in genetic risk for extreme, negative responses to traumatic events. Since it is known that genetic variants are involved in the reporting of events as traumatic and given that trauma is prerequisite for a diagnosis of PTSD, the PTSD risk scores are therefore, in part, representative of individual differences in sensitivity to trauma. UK Biobank participants with higher genetic risk for PTSD may therefore be likely to evaluate events as emotionally distressing and report them accordingly in the Mental Health Questionnaire, compared to individuals with lower genetic risk for PTSD.

This finding is interesting in light of our hypothesis that PTSD would show higher genetic overlap with MDD with reported trauma compared to MDD without reported trauma. Although the genetic correlation analysis yielded no conclusive results, the findings from the PRS analysis provide tentative evidence for an association between the genetics of PTSD and the experience of traumatic life events in those who have MDD. In the genetic correlation analysis, all four PTSD phenotypes (UK Biobank, PGC 1.5 and 2, and the MVP), had greater genetic overlap with MDD with reported trauma compared to MDD without reported trauma. However, the differences between the correlations were not significant. This may be due to the limited power of the original GWASs from which the summary statistics were created. Given the findings from the PRS analysis, it is possible that the greater genetic correlation between PTSD and MDD with reported trauma, compared to MDD without reported trauma, might have been significant if the MDD summary statistics had been produced from larger, better powered GWASs. Therefore, replication with larger MDD GWASs will be useful in understanding whether the differences between the correlations are due to chance. The Genetic Links to Anxiety and Depression (GLAD) study, which aims to recruit 40,000 participants, will provide an opportunity to achieve this with sufficient power in the future.

A further interesting finding from this study is the significantly higher genetic correlation between PTSD and recurrent MDD compared to PTSD and MDD without reported trauma, a finding which was consistent across the UK Biobank and PGC PTSD phenotypes. This finding might reflect similarities in severity between PTSD and recurrent MDD. It is known that exposure to trauma, especially in childhood, is related to MDD that is severe and treatment resistant, as well as recurrent, in later life (Nanni *et al*., 2012; Danese, 2020). Potentially, the MDD without reported trauma phenotype may include participants who have had milder experiences of MDD, while the recurrent MDD phenotype may capture participants with more severe MDD. Like recurrent MDD, PTSD is a severe and disabling psychiatric disorder, where full, clinically significant symptoms can present for months up to years following exposure. It is known that symptoms often persist for years after remission (Kessler *et al*., 2017). Taking this into consideration, PTSD and recurrent MDD might share greater genetic overlap due to similarities in symptom severity and persistence. These similarities may be shared to a lesser extent between PTSD and MDD without reported trauma, which could explain the significant difference between the genetic correlations. Although these conclusions are speculative, an interesting next step would be to calculate the genetic relationship of PTSD with mild and severe MDD in a large cohort with detailed symptom level data on both MDD and PTSD.

### Merits and limitations

We were able to use a variety of PTSD definitions from the largest GWAS of PTSD to date, obtained from samples which recruited participants who were exhibiting differing levels of severity and were recruited in distinct ways. These factors may influence the phenotype’s genetic sharing with MDD. To participate in the UK Biobank, individuals visited recruitment centres for a number of hours to undergo physical assessments, provide data and a DNA sample (Sudlow *et al*., 2015). This level of investment may mean that people who were experiencing severe emotional and functional impairment were unlikely to participate. Contrastingly, the majority of the PGC’s participants were recruited directly from clinically ascertained studies of PTSD, using telephone diagnostic interviews and face-to-face clinical assessments (Nievergelt *et al*., 2019). Consequently, it is reasonable to assume that, on average, the participants comprising the PGC’s data report more severe symptoms than individuals drawn from the population without specific ascertainment for mental ill health (as is the case with the UK Biobank). The benefit of using PTSD phenotypes from samples ascertained using different recruitment methods is that it allowed us to examine how the genetics of MDD differentially relates to PTSD depending on sample-specific features. We saw that the significant difference in genetic correlation between PTSD and recurrent MDD and MDD without reported trauma replicated when using the PGC samples, suggesting this result is not only applicable to UK Biobank participants with PTSD.

Nonetheless, the MDD and trauma-related phenotypes were defined in UK Biobank participants, who show a “volunteer selection bias” (Fry *et al*., 2017) which refers to the tendency of research participants to be more health-conscious and have a higher level of social capital than non-participants (Manolio *et al*., 2012). Furthermore, individuals who completed the follow-up Mental Health Questionnaire, compared with UK Biobank participants overall and the general population, are more likely to have a university degree, come from a higher socioeconomic background and report fewer disabilities and fewer chronic health problems (Davis *et al*., 2019). Therefore, although the UK Biobank offers the opportunity to amalgamate genetic and phenotypic in a large, homogenous, single-population cohort, its demographic features mean the MDD and trauma phenotypes may not be representative of the experiences of wider populations.

In contrast to the UK Biobank and PGC, the MVP sample was limited to United States veterans (Stein *et al*., 2020). Therefore, an interesting finding was that the significant difference in the genetic correlations between PTSD with recurrent MDD and with PTSD and MDD without reported trauma did not replicate when using the MVP PTSD phenotype. There are a number of potential explanations for this. War and combat-related PTSD may be genetically distinct from PTSD arising from other types of catastrophic events. For instance, the World Mental Health Survey showed that war- and combat-related PTSD tends to be longer-lasting than other types (Kessler *et al*., 2017), with a mean symptom duration of 161.7 months for those with combat experience. These factors may alter the way the disorder overlaps genetically with MDD. As shown in Table 1, at least a quarter of the sample had been exposed to combat (Stein *et al*., 2020). Another factor to consider is the demographics of the sample, which overly represents males (94.4%), unlike the UK Biobank, PGC 1.5 and PGC 2 samples, which have an almost even sex division. Previous GWAS findings suggest that the genetics of PTSD may differ between men and women (Nievergelt *et al*., 2019). Therefore, the lack of replication using the PTSD data from the MVP may be attributable to sample-specific characteristics, including sex, severity and trauma type. Overall, using varied PTSD phenotypes in our study has been helpful in understanding how the genetics of differentially relate to MDD.

The interpretation of the results in this study is also affected by limitations of the measure of trauma. Trauma exposure was measured retrospectively, which can lead to inaccurate reporting of events (Colman *et al*., 2016). The relative age of the participants in the UK Biobank (40-69 at baseline) may compromise the accuracy of recall of the events, particularly those in childhood (Table 2). A second issue is the lack of temporal information regarding the onset of MDD in relation to traumatic experiences, which means we cannot infer causality between them. This should be recognised when considering the significantly higher rates of trauma among UK Biobank participants with recurrent MDD compared to single-episode MDD. We cannot assume that this association is causal or that the traumatic events happened before the onset of MDD. To overcome this problem we could have limited the definition of trauma to the three childhood items (Table 2) which would allow more robust measurement of the influence of trauma exposure on the later development of MDD. However, Coleman et al. (2020) reported that limiting the GWAS of MDD with reported trauma to only the events in the Childhood Trauma Screener (Table 2) did not significantly alter the SNP-based heritability of MDD, suggesting that the inclusion of the adulthood events is valid when investigating the relationship between reported trauma and MDD (Coleman *et al*., 2020). Overall, although this method of assessing trauma exposure is not ideal, it is the only feasible method for collecting large amounts of data required for genomic analyses such as those in this study.

Lastly, our results may not generalise to non-European populations, since the GWASs were based on participants of European ancestries only. This limitation, which means the experiences of non-European individuals fail to be accounted for in genetics research, is increasingly being acknowledged. Another recent PTSD GWAS from the MVP also included individuals of African ancestries (Stein *et al*., 2020). Sample sizes are small but will hopefully grow as the field responds to the need for inclusivity and diversity in its research.

### Summary

We emphasise three note-worthy findings. Firstly, individuals in the UK Biobank who have experienced recurrent depressive episodes report significantly higher rates of traumatic life events compared to those who have experienced a single depressive episode. This corroborates previous research which has found that trauma is strongly associated with the development of recurrent MDD (Otte *et al*., 2016; Danese, 2020). Secondly, we report medium to high genetic correlations between PTSD and the four MDD phenotypes (*r*_g_=0.47-0.81). This is consistent with previous genome-wide analyses demonstrating that PTSD and MDD are strongly genetically overlapping. Lastly, we report that higher genetic risk for PTSD is associated with reporting exposure to trauma in individuals with MDD. This could be considered evidence for a heritable basis for trauma sensitivity which influences the way in which a person perceives and responds to traumatic events.

## Data Availability

Genome-wide association study (GWAS) summary statistics for the posttraumatic stress disorder (PTSD) phenotypes were obtained from the PTSD working group of the Psychiatric Genomics Consortium and the Million Veteran Program. GWAS summary statistics for major depressive disorder were obtained from the corresponding author (J.C.) at King's College London. UK Biobank data is available to bona fide researchers with an approved application.

## Data availability

Genome-wide association study (GWAS) summary statistics for the posttraumatic stress disorder (PTSD) phenotypes were obtained from the PTSD working group of the Psychiatric Genomics Consortium and the Million Veteran Program. GWAS summary statistics for major depressive disorder (MDD) were obtained from the corresponding author (J.C.) at King’s College London. UK Biobank data is available to bona fide researchers with an approved application.

## Author contributions

J.M., J.C., and G.B. were responsible for study conception and design. J.C., G.B., M.B.S., The Million Veteran Program and the PTSD working group of the PGC were responsible for acquisition of the data. J.M., and J.C. and M.S. were responsible for data analysis. All authors were involved in the interpretation of the data. J.M. was responsible for drafting of the paper, under the close supervision of J.C. and G.B. All authors read, edited and approved the final manuscript before submission. All authors agree to be accountable for all aspects of the work, and in ensuring that questions related to the accuracy or integrity of any part of the work are appropriately investigated.

## Ethics

The UK Biobank is approved by the North West Multi-centre research Committee. All procedures performed in studies involving human participants were in accordance with the ethical standards of this committee and with the 1964 Declaration of Helsinki and its later amendments or comparable ethics standards. All participants provided written informed consent to participate in the study. This study has been completed under UK Biobank approved study application 16577 (Professor Gerome Breen).

## Acknowledgements

We are indebted to the scientists involved in the construction of the UK Biobank and the PGC, and to the thousands of subjects who have shared their life experiences. This research used data derived from the UK Biobank Resource under application 16577 (Prof Breen). The views expressed are those of the authors and not necessarily those of the National Health Service (NHS), the NIHR or the Department of Health and Social Care.

## Disclosures

The authors declare no conflicts of interest.

## Notes

### Competing Interest Statement

The authors have declared no competing interest.

### Funding Statement

This study represents research funded partly by the National Institute for Health Research (NIHR) Maudsley Biomedical Research Centre at South London and Maudsley NHS Foundation Trust and King's College London, and partly by the Lord Leverhulme Charitable grant (Professor Robin Murray). High performance computing facilities were funded with capital equipment grants from the GSTT Charity (TR130505) and Maudsley Charity (980).

### Summary of Updates

I added an author to the author list. This author had mistakenly been missed off during the first upload.

