## Supplementary material for "Psychological trauma and the genetic overlap between posttraumatic stress disorder and major depressive disorder": Supplementary_material_tables_JM.pdf

### <u>Methods</u>

#### Contributing studies and phenotype definitions

##### *UK Biobank*

Between 2006 and 2010, the UK Biobank recruited 502,617 individuals aged 40-69 for a cohort study aimed at improving diagnosis, treatment and prevention of serious diseases (Allen *et al.*, 2014). Participants gave full informed consent, answered surveys and provided physical measurements including DNA samples at a baseline visit to one of 22 assessment centres across the UK (Sudlow *et al.*, 2015). The phenotypes assessed in this paper were derived from the online follow-up Mental Health Questionnaire (MHQ), which received 157,358 responses. This online questionnaire comprises a number of adapted versions of clinically used psychiatric questionnaires to assess common mental health disorders. Case definitions based on responses to the psychiatric questionnaires were derived by the working committee who wrote the MHQ (Davis *et al.*, 2019).

*Posttraumatic stress disorder phenotype*

The definition of PTSD in the UK Biobank is included in the Supplementary Material of Nievergelt et al. (2019). In brief, the PTSD phenotype was derived from six questions asked in the follow-up online mental health questionnaire. These questions were derived from the brief civilian version of the PTSD Checklist Screener (PCL-S) which measures PTSD symptoms experienced in the previous month: avoidance of activities; disturbing thoughts; and feeling upset; and two additional questions related to feeling distant and feeling irritable. Each item was scored on a five-point Likert item measuring the amount of concern caused by that symptom in the past month (1="Not at all" to 5="Extremely"). In addition, a “trouble concentrating” question from the Patient Health Questionnaire-9 (PHQ9) depression questionnaire was added to replace a similar item that would normally be included in the PCL-S. This item was scored on a four-point Likert item according to frequency of difficulties associated with trouble concentrating (1="Not at all" and 4="Nearly every day"). For each participant, all items were summed into a total score ranging 3-29. Participants were considered PTSD cases if they had an overall PCL-S score ≥ 13. Participants were considered PTSD controls if they responded to all of the initial three questions and had PCL-S score ≤ 12 (Nievergelt *et al.*, 2019).

*Major depressive disorder phenotype*

Phenotype definitions for “MDD with reported exposure to psychological trauma” and “MDD without reported exposure to psychological trauma” are included in the Supplementary Material of Coleman et al, 2020. Participants were considered cases for probable MDD based on their responses to questions derived from the Composite International Diagnostic Interview Short Form (CIDI-SF). Reporting on a period of depression lasting at least two weeks, cases endorsed at least one of the two core symptoms (“ever had prolonged feelings of sadness or depression” and “ever had prolonged loss of interest in normal activities”), at least five of the nine symptoms queried overall, and reported that they were affected almost every day, most days during the period, with more than a little impact on normal functioning. Controls did not meet case criteria and did not meet criteria for a current episode of depression. Participants who self-reported a diagnosis of schizophrenia, other psychoses, or bipolar disorder were excluded. Controls were excluded if they self-reported any mental illness, taking any drug with an antidepressant indication, or had been hospitalised with a mood disorder or met previously defined criteria for a mood disorder.

*MDD and the presence/absence of psychological trauma exposure phenotypes*

Questions relating to traumatic experiences in childhood were assessed on a five point scale (ranging from never to often) using the Childhood Trauma Screener (an adapted version of the Childhood Trauma Questionnaire (Bernstein *et al.*, 1994; Grabe *et al.*, 2012; Bellis *et al.*, 2014). An equivalent screener was constructed for traumatic events in adulthood. Only traumatic experiences with an odds ratio >2.5 with MDD were selected to obtain a single binary variable for trauma exposure. This included: three events in childhood (did not feel loved, felt hated by a family member, sexually abused); three events in adulthood (physical violence, belittlement, sexual interference); and one PTSD-relevant event (ever a victim of sexual assault). Participants were included in the "MDD with reported exposure to psychological trauma" analysis if they reported two or more of these events and met case or control criteria for MDD. Participants were included in the "MDD without reported exposure to psychological trauma" analysis if they reported none of these events and met case or control criteria for MDD (Coleman *et al.*, 2020).

*Recurrent and single-episode major depressive disorder phenotypes*

The further definitions of recurrent and single-episode MDD can be found in the Supplementary Material of Coleman et al. (2019). In brief, participants who met criteria for MDD were classified with recurrent MDD if they reported multiple depressed periods across their lifetime and single-episode MDD otherwise (Coleman *et al.*, 2019).

***Posttraumatic Stress Disorder working group of the Psychiatric Genomics Consortium***

The PTSD working group of the Psychiatric Genomics Consortium (PGC) meta-analysed data from 59 studies of PTSD to perform a GWAS known as the PGC-PTSD 1.5. To maximise power in genome-wide analysis, the UK Biobank and PTSD Freeze 1.5 (PGC 1.5 PTSD) were combined. This combined data set is known as the PTSD Freeze 2 (PGC 2 PTSD).

*Posttraumatic stress disorder phenotypes*

The PGC gathered data for the PTSD Freeze 1.5 through a number of independent studies who used a wide range of methods, primarily telephone diagnostic interviews and face-to-face clinical assessments. Many participants included in this cohort are veterans who have been combat or war-zone exposed. Other traumatic events assessed by the PGC include serious car accidents, campus shootings, domestic violence, and childhood physical and sexual abuse. Participants were assessed for current and lifetime PTSD using various instruments and different versions of the DSM (Nievergelt *et al.*, 2019). Further details of the contributing studies and instruments used to assess PTSD are contained in the Supplementary Material of Nievergelt et al. (2019).

#### ***The Million Veteran Program***

The Million Veteran Program (MVP) is a longitudinal study of United States military veterans who provided a blood sample for biobanking and responses to various questionnaires (Gaziano *et al.*, 2016).

*Posttraumatic stress disorder phenotype*

The MVP performed GWAS on an algorithmically-defined probable PTSD phenotype. While the MVP sample consists of United States veterans, only 27.5% have confirmed war- or combat- exposure, while 29.3% who had not been exposed. The remaining 43.1% had unknown war- and combat-exposure (Stein *et al.*, 2020). Further details of how the two MVP phenotypes were defined can be found in Stein et al. (2020). The MVP PTSD GWAS was of a binary phenotype based on the veterans' Electronic Health Records.

### Computational Methods

#### ***High Definition Likelihood inference of genetic correlations***

Genetic correlations were estimated using High Definition Likelihood (HDL). Firstly for each phenotype, GWAS summary statistics were used to estimate the proportion of variance explained by common genetic variants ($h^2_{SNP}$) using High Definition Likelihood (HDL). While this is not a specific aim of the study, this step is necessary for interpreting genetic correlations between the four PTSD phenotypes and the four MDD-related phenotypes. HDL rests upon the principle of linkage-disequilibrium and extends the regression formula used by Linkage Disequilibrium Score Regression (LDSC) (see below). Unlike LDSC, HDL uses a full, likelihood-based method to estimate genetic correlations (Ning *et al.*, 2020). Further details can be found in the original paper by Ning et al. (2020).

All summary statistics were wrangled using the built-in HDL function for data wrangling (https://github.com/zhenin/HDL/wiki/Format-of-summary-statistics). All GWAS summary statistics used in this analysis had at least 99% SNP overlap with the UK Biobank LD reference panel using HapMap3 variants, apart from the Million Veteran Program (MVP) GWAS which had an overlap of 94.23%. Through correspondence with the HDL authors, we are confident that this level of SNP overlap is acceptable since the mismatch is not due to differences in the ancestral population from which the samples were created (e.g European vs. non-European populations - both the MVP and the UK Biobank samples were made up of participants from European ancestries). The authors of HDL have performed a simulation to test this and discovered that missing SNPs lead to more conservative results but should not generate false positives (Z Ning, personal correspondence and authorisation by email). Therefore, a missing rate around 5% is acceptable.

Furthermore, we repeated the data wrangling using the smaller HapMap2 (as opposed to HapMap3) reference panel and this did not improve the SNP overlap appreciably (data not shown). The authors of HDL therefore advised us that we continue to use the

HapMap3 reference panel for the wrangling of the MVP Case-Control GWAS summary statistics.

***High Definition Likelihood block-jackknife***

The block-jackknife method was used to compare genetic correlations for a statistically significant difference between them. Each genetic correlation was compared in a pairwise fashion with all other genetic correlations within each set (the sets being UK Biobank PTSD, PGC 1.5 PTSD, PGC 2 PTSD, and MVP Case-Control PTSD). The block-jackknife uses a resampling method to estimate standard errors for each genetic correlation, which is then used to determine whether the differences between the genetic correlations are significantly different to zero. Here we provide an explanation of the block-jackknife method (Z Ning, personal correspondence and authorisation by email).

When estimating genetic correlations, $r_g1$ and $r_g2$, you may then want to test whether the difference between them, referred to as the "global difference", is significantly different to zero. This would mean our null hypothesis is: $r_g1 - r_g2 = 0$. For $r_g1$, by setting jackknife.df=TRUE when using the HDL tool, you can get jackknife estimates of $r_g1.1$ to $r_g1.61$. This creates a file with 61 jackknife estimates because the genome is split into 61 pieces during the resampling process. $r_g1.k$ represents the estimated $r_g$ with piece k removed. Similarly, you have $r_g2.1$ to $r_g2.61$ for $r_g2$.

Once you have two files with these values in for both correlations, you can create the block-jackknife estimates by doing $r_g1.1 - r_g2.1$ all the way up to $r_g1.61 - r_g2.61$. You can then get the block-jackknife standard error using this formula:

$$\widehat{se_{Jack}} = \sqrt{\frac{n-1}{n}\sum_{i=1}^{n}\left(\hat{\theta}_{(i)} - \bar{\hat{\theta}}_{(.)}\right)^2}$$

Where $n$ = 61 (for each of the resampling estimates), $\hat{\theta}$ = the difference between each resampling estimate (e.g. $r_g1.1$ - $r_g2.1$ all the way up to $r_g1.61$ - $r_g2.61$), $\bar{\hat{\theta}}$ = the mean of these estimates.

Once you have calculated the block-jackknife standard error, you can use this and the global difference between the two correlations to do a Wald test:

Z = global difference ÷ block-jackknife standard error

The Wald test will give you a z-score, which you can then use to find the two-tailed p-value, which will tell you whether the global difference is significantly different to zero. In our study, we corrected for multiple testing by considering $p<0.008$ as the threshold for significance (0.05/6 = 0.008, to account for the 6 block-jackknife tests carried out for each PTSD phenotype). Supplementary Table 1 contains all the results from the HDL block-jackknife analysis.

**<u>Converting observed scale heritability estimates to the liability scale</u>**

The heritability of each trait was estimated by the HDL programme irrespective of population prevalence. Therefore, these estimates were converted to the liability scale in R, using code from the Nievergelt Lab github: https://gist.github.com/nievergeltlab/fb8a20feded72030907a9b4e81d1c6ea. Standard errors were also converted to the liability scale using the same formula (Lee *et al.*, 2011). Population prevalence for PTSD was adopted from Nievergelt et al. (2019), from Coleman et al. (2020) for reported trauma in MDD and from Burcusa & Iacono (2007) for recurrence in MDD.

***Linkage Disequilibrium Score (LDSC) Regression***

In addition to using HDL, we calculated genetic correlations using Linkage Disequilibrium Score (LDSC) Regression. LDSC Regression is a command line tool for estimating heritability and genetic correlation from GWAS summary statistics. It rests upon the principle of linkage-disequilibrium (LD). LD describes the degree to which an allele of one SNP is inherited or correlated with an allele of another SNP within a population(Bush and Moore, 2012). In this method, an “LD Score” of a given SNP refers to “the sum of LD $r^2$ measured with all other SNPs”. LDSC works by performing regression analysis on the LD scores and the test statistic of each SNP included in the GWAS, including those that do not meet genome-wide significance(Bulik-Sullivan *et al.*, 2015a). LDSC relies on the fact that the GWAS effect-size estimate for a given SNP incorporates the effect of all SNPs in LD with that particular SNP. For most complex human traits, which are polygenic, SNPs with high LD will have higher chi-square test statistics, on average, than SNPs with low LD (Bulik-Sullivan *et al.*, 2015b).

As in the HDL analysis, GWAS summary statistics were used to estimate the proportion of variance explained by common genetic variants ($h^2_{SNP}$) using LDSC Regression. While this is not a specific aim of the study, this step is necessary for interpreting genetic correlations.

#### *Linkage Disequilibrium Score (LDSC) block-jackknife*

The block-jackknife method was used to compare genetic correlations for a statistically significant difference between them. Each genetic correlation was compared in a pairwise fashion with all other genetic correlations within each set (the sets being UK Biobank PTSD, PGC 1.5 PTSD, PGC 2 PTSD, and MVP Case-Control PTSD). As described above for HDL, the block-jacknife works by repeated re-estimation of blocks of jack-knife estimates. In the case of LDSC, the number of blocks is set by the user (we used 200), but otherwise the calculation of significant differences follows the description provided for HDL.

#### *Polygenic Risk Scores*

PRS were calculated PRSice v2.3.1, a command line programme that uses GWAS summary statistics to calculate genetic risk of a base phenotype in individuals from an independent sample. A PRS refers to the summation of alleles across many genetic loci associated with a particular trait or disease. These alleles are typically weighted by effect sizes estimated from GWAS (Euesden *et al.*, 2015). In our study, the PRS represents the aggregated PTSD risk conferred by many DNA variants in participants of the UK Biobank who have MDD. Since even well-powered GWAS offer only tentative evidence for causally associated variants, PRS are calculated at a range of different *P*-value thresholds to provide the 'best-fit', or most predictive, PRS (Dudbridge, 2013). Once the best-fit PRS has been estimated, these are used as predictors of a target phenotype in individuals in an independent sample in a regression. PRS were calculated at 11 p-value thresholds ($5x10^{-8}$, $1x10^{-5}$, $1x10^{-3}$, 0.01, 0.05, 0.1, 0.2, 0.3, 0.4, 0.5, 1). Phenotype permutations (implemented in PRSice) were used to produce an empirical p-value for the association at the best-fitting PRS, which accounts for testing multiple thresholds. We then performed a logistic regression to examine whether the risk scores are more strongly associated with MDD with reported trauma or MDD without reported trauma, and recurrent or single-episode MDD.

***Power calculations***

We calculated the power of our PRS analyses using the Additive Variance Explained and Number of Genetic Effects Method of Estimation (AVENGEME) programme in R (Dudbridge, 2013). We were uncertain about the covariance between genetic effects in the target sample (MVP PTSD) and the two target samples. Therefore, based on the parameters required by the AVENGEME package (below), we calculated that for the PRS analysis testing the risk score's association with MDD with and without reported trauma to be powered at at least 80%, the covariance between the genetic effect sizes in the training and target samples would need to be at least 0.024. We calculated that for the

PRS analysis testing the risk score's association with recurrent and single-episode MDD to be powered at at least 80%, the covariance between the genetic effect sizes in the training and target samples would need to be at least 0.0305. Due to these minimum covariance estimates being so low, we were confident that the PRS analyses were powered by at least 80%.

*Parameters required by the AVENGEME package:*

- Number of SNPs included in PRS after clumping = 183,881
- Proportion of variance in PTSD explained by genetic effects = 0.03 (Stein *et al.*, 2020)
- Training sample prevalence of PTSD = 0.18 (Stein *et al.*, 2020)
- Population prevalence of PTSD = 0.30 (Nievergelt *et al.*, 2019)
- Target sample prevalence of reported trauma among MDD cases = 0.59
- Population prevalence of reported trauma among MDD cases = 0.52 (Coleman *et al.*, 2020)
- Target sample prevalence of recurrent MDD = 0.59
- Population prevalence of recurrent MDD among MDD cases = 0.5 (Burcusa and Iacono, 2007)

### Results

#### High Definition Likelihood block-jackknife

The results of the HDL block-jackknife analysis can be found in Supplementary Table 1. Differences between genetic correlations were considered statistically significant if they surpassed the Bonferroni corrected alpha (0.05/6 = 0.008; i.e. to correct for the six block-jackknife tests per PTSD phenotype).

Supplementary Table 1: genetic correlation results from the High Definition Likelihood (HDL) block-jackknife analysis of posttraumatic stress disorder (PTSD) and the four major depressive disorder (MDD) phenotypes. Each genetic correlation was compared in a pairwise fashion with all other genetic correlations in the set (i.e. for each PTSD phenotype). Differences between genetic correlations were considered statistically significant if they passed the Bonferroni adjusted threshold ($p<0.008$) are shown in bold. PGC refers to Psychiatric Genomics Consortium and MVP refers to Million Veteran Program.

| HDL genetic correlation 1 | HDL genetic correlation 2 | $r_g$ difference | SE | P (diff 0) |
|---|---|---|---|---|
| UK Biobank PTSD and MDD with reported trauma | UK Biobank PTSD and MDD without reported trauma | 0.1339 | 0.09 | 0.14 |
| UK Biobank PTSD and MDD with reported trauma | UK Biobank PTSD and recurrent MDD | -0.1094 | 0.06 | 0.06 |
| UK Biobank PTSD and MDD with reported trauma | UK Biobank PTSD and single-episode MDD | -0.0426 | 0.09 | 0.62 |
| UK Biobank PTSD and MDD without reported trauma | UK Biobank PTSD and recurrent MDD | -0.2433 | 0.07 | **$4.77 \times 10^{-4}$** |
| UK Biobank PTSD and MDD without reported trauma | UK Biobank PTSD and single-episode MDD | -0.1765 | 0.08 | 0.02 |
| UK Biobank PTSD and recurrent MDD | UK Biobank PTSD and single-episode MDD | 0.0668 | 0.08 | 0.37 |
| PGC 1.5 PTSD and MDD with reported trauma | PGC 1.5 PTSD and MDD without reported trauma | 0.0679 | 0.10 | 0.49 |
| PGC 1.5 PTSD and MDD with reported trauma | PGC 1.5 PTSD and recurrent MDD | -0.1417 | 0.07 | 0.05 |
| PGC 1.5 PTSD and MDD with reported trauma | PGC 1.5 PTSD and single-episode MDD | -0.2040 | 0.14 | 0.15 |
| PGC 1.5 PTSD and MDD without reported trauma | PGC 1.5 PTSD and recurrent MDD | -0.2096 | 0.07 | **$4.90 \times 10^{-3}$** |
| PGC 1.5 PTSD and MDD without reported trauma | PGC 1.5 PTSD and single-episode MDD | -0.2719 | 0.12 | 0.02 |
| PGC 1.5 PTSD and recurrent MDD | PGC 1.5 PTSD and single-episode MDD | -0.0623 | 0.12 | 0.61 |
| PGC 2 PTSD and MDD with reported trauma | PGC 2 PTSD and MDD without reported trauma | 0.0988 | 0.13 | 0.43 |
| PGC 2 PTSD and MDD with reported trauma | PGC 2 PTSD and recurrent MDD | -0.1418 | 0.09 | 0.10 |
| PGC 2 PTSD and MDD with reported trauma | PGC 2 PTSD and single-episode MDD | -0.1650 | 0.10 | 0.12 |
| PGC 2 PTSD and MDD without reported trauma | PGC 2 PTSD and recurrent MDD | -0.2406 | 0.09 | **$5.28 \times 10^{-3}$** |

| | | | | |
|---|---|---|---|---|
| PGC 2 PTSD and MDD without reported trauma | PGC 2 PTSD and single-episode MDD | -0.2638 | 0.11 | 0.01 |
| PGC 2 PTSD and recurrent MDD | PGC 2 PTSD and single-episode MDD | 0.0232 | 0.10 | 0.82 |
| MVP PTSD and MDD with reported trauma | MVP PTSD and MDD without reported trauma | 0.0538 | 0.12 | 0.65 |
| MVP PTSD and MDD with reported trauma | MVP PTSD and recurrent MDD | -0.0203 | 0.07 | 0.77 |
| MVP PTSD and MDD with reported trauma | MVP PTSD and single-episode MDD | -0.0894 | 0.10 | 0.38 |
| MVP PTSD and MDD without reported trauma | MVP PTSD and recurrent MDD | -0.0741 | 0.09 | 0.40 |
| MVP PTSD and MDD without reported trauma | MVP PTSD and single-episode MDD | -0.1432 | 0.11 | 0.18 |
| MVP PTSD and recurrent MDD | MVP PTSD and single-episode MDD | -0.0691 | 0.11 | 0.54 |

**LDSC Regression genetic correlations**

Genetic correlations (Supplementary Table 2) estimated by LDSC Regression were considered significantly different to zero and to one if they reached or surpassed the Bonferroni-corrected alpha in each analysis (0.05/4 = 0.0125; i.e. to correct for the four tests).

Supplementary Table 2: Linkage Disequilibrium Score (LDSC) Regression genetic correlation estimates ($r_g$) and standard errors (SE) of 1) UK Biobank posttraumatic stress disorder (PTSD), 2) Psychiatric Genomics Consortium (PGC) 1.5 PTSD, 3) PGC 2 PTSD, 4) Million Veteran Program PTSD with the four major depressive disorder (MDD)-related phenotypes. *P* (diff 0) refers to *p*-value for test of $r_g$ different from 0. *P* (diff 1) refers to *p*-value for test of $r_g$ different from 1. Genetic correlations were considered significant if they passed the Bonferroni adjusted threshold ($p<0.0125$).

| PTSD Phenotype | MDD phenotype | $r_g$ | SE | P (diff 0) | P (diff 1) |
|---|---|---|---|---|---|
| **UK Biobank PTSD** | MDD with reported trauma | 0.6499 | 0.08 | $5.70 \times 10^{-15}$ | $2.58 \times 10^{-5}$ |
| **UK Biobank PTSD** | MDD without reported trauma | 0.6538 | 0.14 | $1.67 \times 10^{-6}$ | 0.01 |
| **UK Biobank PTSD** | Recurrent MDD | 0.7815 | 0.06 | $7.55 \times 10^{-46}$ | $7.12 \times 10^{-5}$ |
| **UK Biobank PTSD** | Single-episode MDD | 0.7158 | 0.10 | $7.29 \times 10^{-12}$ | $6.54 \times 10^{-3}$ |
| **PGC 1.5 PTSD** | MDD with reported trauma | 0.6976 | 0.21 | $1.06 \times 10^{-3}$ | 0.16 |
| **PGC 1.5 PTSD** | MDD without reported trauma | 0.4514 | 0.25 | $6.77 \times 10^{-2}$ | 0.03 |
| **PGC 1.5 PTSD** | Recurrent MDD | 0.7438 | 0.15 | $1.48 \times 10^{-6}$ | 0.10 |
| **PGC 1.5 PTSD** | Single-episode MDD | 0.5743 | 0.17 | $7.07 \times 10^{-4}$ | 0.01 |

| | | | | | |
|---|---|---|---|---|---|
| **PGC 2 PTSD** | MDD with reported trauma | 0.6263 | 0.12 | $6.72 \times 10^{-8}$ | $1.27 \times 10^{-3}$ |
| **PGC 2 PTSD** | MDD without reported trauma | 0.5461 | 0.17 | $1.15 \times 10^{-3}$ | $6.90 \times 10^{-3}$ |
| **PGC 2 PTSD** | Recurrent MDD | 0.7592 | 0.09 | $6.47 \times 10^{-18}$ | $6.21 \times 10^{-3}$ |
| **PGC 2 PTSD** | Single-episode MDD | 0.6597 | 0.12 | $1.32 \times 10^{-8}$ | $3.38 \times 10^{-3}$ |
| **MVP PTSD** | MDD with reported trauma | 0.3906 | 0.08 | $3.40 \times 10^{-7}$ | $1.78 \times 10^{-15}$ |
| **MVP PTSD** | MDD without reported trauma | 0.3593 | 0.10 | $5.15 \times 10^{-4}$ | $6.00 \times 10^{-10}$ |
| **MVP PTSD** | Recurrent MDD | 0.4812 | 0.06 | $1.30 \times 10^{-14}$ | $9.24 \times 10^{-17}$ |
| **MVP PTSD** | Single-episode MDD | 0.4392 | 0.09 | $6.84 \times 10^{-7}$ | $2.35 \times 10^{-10}$ |

**Linkage Disequilibrium Score Regression block-jackknife**

We tested the differences between the genetic correlations using a block-jackknife. Differences were considered statistically significant if they passed a Bonferroni-corrected alpha of 0.008 (0.05/6 = 0.008; i.e. to correct for the six block-jackknife tests per PTSD phenotype). Supplementary Table 3 contains the results of the LDSC Regression block-jackknife.

Supplementary Table 3: genetic correlation results from the Linkage Disequilibrium Score Regression (LDSC) block-jackknife analysis of posttraumatic stress disorder (PTSD) and the four major depressive disorder (MDD) phenotypes. Each genetic correlation was compared in a pairwise fashion with all other genetic correlations in that set. Differences between genetic correlations were considered statistically significant if they passed the Bonferroni adjusted threshold ($p<0.008$) are shown in bold. PGC refers to Psychiatric Genomics Consortium and MVP refers to Million Veteran Program.

| LDSC genetic correlation 1 | LDSC genetic correlation 2 | $r_g$ difference | SE | P (diff 0) |
|---|---|---|---|---|
| UK Biobank PTSD and MDD with reported trauma | UK Biobank PTSD and MDD without reported trauma | -0.0039 | 0.17 | 0.99 |
| UK Biobank PTSD and MDD with reported trauma | UK Biobank PTSD and recurrent MDD | -0.1316 | 0.08 | 0.08 |
| UK Biobank PTSD and MDD with reported trauma | UK Biobank PTSD and single-episode MDD | -0.0659 | 0.12 | 0.55 |
| UK Biobank PTSD and MDD without reported trauma | UK Biobank PTSD and recurrent MDD | -0.1277 | 0.12 | 0.25 |
| UK Biobank PTSD and MDD without reported trauma | UK Biobank PTSD and single-episode MDD | -0.0620 | 0.13 | 0.57 |
| UK Biobank PTSD and recurrent MDD | UK Biobank PTSD and single-episode MDD | 0.0657 | 0.11 | 0.51 |
| PGC 1.5 PTSD and MDD with reported trauma | PGC 1.5 PTSD and MDD without reported trauma | 0.2462 | 0.29 | 0.40 |
| PGC 1.5 PTSD and MDD with reported trauma | PGC 1.5 PTSD and recurrent MDD | -0.0462 | 0.17 | 0.76 |
| PGC 1.5 PTSD and MDD with reported trauma | PGC 1.5 PTSD and single-episode MDD | 0.1233 | 0.20 | 0.58 |
| PGC 1.5 PTSD and MDD without reported trauma | PGC 1.5 PTSD and recurrent MDD | -0.2924 | 0.21 | 0.16 |
| PGC 1.5 PTSD and MDD without reported trauma | PGC 1.5 PTSD and single-episode MDD | -0.1229 | 0.23 | 0.57 |
| PGC 1.5 PTSD and recurrent MDD | PGC 1.5 PTSD and single-episode MDD | 0.1695 | 0.18 | 0.36 |
| PGC 2 PTSD and MDD with reported trauma | PGC 2 PTSD and MDD without reported trauma | 0.0802 | 0.20 | 0.65 |
| PGC 2 PTSD and MDD with reported trauma | PGC 2 PTSD and recurrent MDD | -0.1329 | 0.11 | 0.23 |
| PGC 2 PTSD and MDD with reported trauma | PGC 2 PTSD and single-episode MDD | -0.0334 | 0.13 | 0.87 |
| PGC 2 PTSD and MDD without reported trauma | PGC 2 PTSD and recurrent MDD | -0.2131 | 0.14 | 0.12 |
| PGC 2 PTSD and MDD without reported trauma | PGC 2 PTSD and single-episode MDD | -0.1136 | 0.16 | 0.49 |
| PGC 2 PTSD and recurrent MDD | PGC 2 PTSD and single-episode MDD | 0.0995 | 0.12 | 0.36 |
| MVP PTSD and MDD with reported trauma | MVP PTSD and MDD without reported trauma | 0.0313 | 0.13 | 0.82 |
| MVP PTSD and MDD with reported trauma | MVP PTSD and recurrent MDD | -0.0906 | 0.07 | 0.14 |
| MVP PTSD and MDD with reported trauma | MVP PTSD and single-episode MDD | -0.0486 | 0.10 | 0.60 |
| MVP PTSD and MDD without reported trauma | MVP PTSD and recurrent MDD | -0.1219 | 0.10 | 0.15 |
| MVP PTSD and MDD without reported trauma | MVP PTSD and single-episode MDD | -0.0799 | 0.10 | 0.40 |
| MVP PTSD and recurrent MDD | MVP PTSD and single-episode MDD | 0.0420 | 0.09 | 0.52 |

314

### Comparison of the two methods for estimating genetic correlations

In our study, we performed the genetic correlation and block-jackknife analyses in both HDL and LDSC Regression. We have opted to only present the HDL results in the main text since it is the preferable method (for reasons discussed in the paper). Nonetheless, there are some key differences between the HDL and LDSC Regression results which need to be discussed here.

An anticipated difference was that no genetic correlations were found to differ significantly from any other genetic correlations when using LDSC Regression. The absence of statistically significant differences in genetic correlation must be interpreted in the context of the power of the original GWAS from which the summary statistics used in this study were created. As shown in Supplementary Table 2, the standard errors surrounding the LDSC Regression point estimates are notably large, meaning that any differences between genetic correlation would need to be large to be detected as significantly different to zero in the block-jackknife analysis. The advantage of using HDL to estimate genetic correlations is reduction in variance of the point estimate (Ning *et al.*, 2020), which provides better power to observe differences between genetic correlations. This was the case in our study, where we found that PTSD is significantly more genetically correlated with recurrent MDD than it is genetically correlated with MDD without reported trauma when using HDL. Similarly to the HDL results, when using LDSC Regression we find that all PTSD phenotypes have a higher genetic correlation with recurrent MDD than with MDD without reported trauma; however, unlike when using the HDL block-jackknife, this difference was not significantly different to zero in the results of LDSC Regression block-jackknife.

Another difference between the HDL and LDSC Regression results is the pattern of results. Firstly, when using LDSC Regression, we observe a clear pattern where all PTSD phenotypes are more genetically correlated with recurrent MDD than single-episode MDD. We do not see this pattern when using HDL. Secondly, the consistent pattern whereby all PTSD phenotypes are more genetically correlated with MDD with reported trauma compared MDD without reported trauma when using HDL only holds true for the PGC 1.5, PGC 2 and MVP PTSD phenotypes when using LDSC Regression, whereas the UK Biobank PTSD phenotype appears to be slightly more genetically correlated with MDD with without reported trauma ($r_g$ difference = 0.0039).

These differences are likely due to inconsistent SNP reference panels used by HDL and LDSC Regression, which are needed to estimate the LD-scores. Therefore, a difference in the reference panel between the two methods is likely to lead to differing results. In HDL, the reference panels with imputed SNPs are based on genotypes in UK Biobank, which were imputed to HRC and UK10K + 1000 Genomes. Specifically in our study, we used the 1,029,876 Quality Controlled UK Biobank imputed HapMap3 SNPs reference panel (https://github.com/zhenin/HDL/wiki/Reference-panels). On the other hand, LDSC Regression uses a reference panel from the 1000 Genomes Project which is not specific to the UK Biobank (Bulik-Sullivan *et al.*, 2015b). According to correspondence with the authors, HDL uses the UK Biobank reference panels instead of 1000 Genomes because the UK Biobank has a larger sample size. This therefore leads to more accurate estimates of LD. Further details can be found at the discussion section of Ning et al. (2020).

However, this increase in the accuracy of LD estimation is likely to apply most strongly to summary statistics from the UK in general and UK Biobank in particular, and less so to samples descended from European ancestry populations from elsewhere in Europe. This being the case, it is feasible that the differences in genetic correlations we observe in our study partly reflect differences in the proportion of UK ancestry in the PTSD summary statistics (as all of the MDD summary statistics were drawn from UK Biobank). This inconsistency between LDSC and HDL, and its potential relationship to UK ancestry in the summary statistics assessed, is likely to have wider implications than our study alone and requires a detailed examination beyond the scope of this study.

**Polygenic risk scores**

Polygenic risk scores for PTSD were calculated PRSice v2.3.1. Two PRS analyses were performed: two regressions using PRS based on the MVP PTSD summary statistics. Full results of these analyses can be found in Supplementary Table 4. Beta coefficients were exponentiated in R to give odds ratios (OR) and 95% confidence intervals (CI).

Supplementary Table 4: Results table from PRSice analysis. Polygenic Risk Scores (PRS) for PTSD were calculated based on the Million Veteran Program (MVP) summary statistics. Table includes the best fitting *p*-value threshold used in the analyses (Threshold), the standardised beta coefficient from regression (Standardised beta) and standard error (SE), the odds ratio (OR) and 95% confidence intervals (CI), *p*-value from regression (*P*), the number of single nucleotide polymorphisms used in creating the risk scores (Number of SNPs) and empirical *p*-value accounting for testing at multiple thresholds (Empirical P). Regression coefficients were considered statistically significant if they surpassed the Bonferroni adjusted alpha ($p<0.025$).

| **Regression** | **Threshold** | **Standardised beta** | **SE** | **OR (95% CI)** | ***P*** | **Number of SNPs** | **Empirical *P*** |
|---|---|---|---|---|---|---|---|
| **MDD with reported trauma vs. MDD without reported trauma** | 0.4 | 0.057 | 0.013 | 1.06 (1.03 – 1.09) | 2.11x$10^{-5}$ | 101111 | 1.0x$10^{-2}$ |
| **Recurrent MDD vs. single-episode MDD** | 0.4 | 0.023 | 0.012 | 1.02 (1.00 – 1.05) | 0.05 | 101111 | 0.28 |

***Nagelkerke's R2***

The PRS were initially calculated without consideration of the prevalence of the target phenotype (reported trauma in MDD and recurrence in MDD) within the population. Based on the results from PRSice, Nagelkerke's R2 was calculated for the estimated population prevalence, +10% and -10% in R. We obtained an estimated population prevalence for trauma exposure among MDD cases of 52% from the Supplementary Material of Coleman et al. (2020). We obtained an estimated population prevalence for recurrence among MDD cases of 50% from Burcusa & Iacono (2007). MDD with reported trauma and MDD without reported trauma, the Nagelkerke's R2 based on a population prevalence of 42%, 52% and 62% were 0.13%, 0.12% and 0.12% respectively. For the regression testing the PTSD risk score's association with recurrent MDD compared to single-episode MDD, the Nagelkerke's R2 based on population prevalence 40%, 50% and 60% was 0.03%, 0.02% and 0.02% respectively.
